# Perinatal Granulopoiesis and Risk of Pediatric Asthma

**DOI:** 10.1101/2020.06.17.20127407

**Authors:** Benjamin A. Turturice, Juliana Theorell, Mary Dawn Koenig, Lisa Tussing-Humphreys, Diane R. Gold, Augusto A. Litonjua, Emily Oken, Sheryl L. Rifas-Shiman, David L. Perkins, Patricia W. Finn

## Abstract

Many perinatal characteristics are associated with risk for pediatric asthma. Identification of biologic processes influenced by these characteristics could facilitate risk stratification or new therapeutic targets. Using publicly available transcriptomic data from CBMCs, transcription of genes involved in myeloid differentiation were inversely associated with pediatric asthma risk stratification based on gestational age at birth, sex, birthweight, and maternal pre-pregnancy BMI. This gene signature was validated in an independent cohort and was specifically associated with genes localizing to neutrophil specific granules. Changes in these genes correlated with changes in protein abundance in serum. CBMC serum levels of PGLYRP-1, a specific granule protein, and sIL6Rα, a membrane protein, were tested for association with pulmonary outcomes. PGLYRP-1 concentration was inversely associated with mid-childhood current asthma and early-teen FEV_1_/FVC×100. Thus, variation in neutrophil specific granule abundance at birth is associated with individual risk for pediatric asthma and reduced pulmonary function in adolescence.

## Introduction

Several risk factors for pediatric asthma can be ascertained in the perinatal period. These risk factors include maternal characteristics (e.g., maternal atopy, maternal BMI, race/ethnicity), demographics (e.g., newborn sex), and birth characteristics (e.g., birthweight, gestational age at birth, mode of delivery) (1). Meta-analyses have provided strong evidence for associations between the variables stated above and risk for pediatric asthma (2-5). Many of these risk factors co-occur (e.g., low birthweight and preterm birth) and it has yet to be discerned whether their imparted risk is mediated through similar biologic processes.

The maternal and fetal immune systems during pregnancy are biased towards Th2 responses and exhibit immune-tolerance to foreign antigens (6-8). Throughout the majority of gestation, fetal hematopoiesis generates mainly lymphoid and erythroid lineages. Towards term gestation, there is a shift towards generation of cells in the myeloid lineages (e.g. neutrophils, monocytes) (9). This phenomenon is evident clinically, as pre-term (<37 weeks gestation) infants have higher lymphocyte to neutrophil ratios compared to those born term (≥37 weeks gestation) (10). The myeloid compartment produces several cytokines (including IL1β, IL-6, IL-18) and co-stimulatory molecules that shift CD4^+^ T-cells away from Th2 responses (11-13). With regards to outcomes, enhanced production of IFNγ by leukocyte stimulation early in life is associated with reduced susceptibility to infections in the lower respiratory tract and recurrent wheezing; however, these findings have not been shown to extend to asthma diagnosis in childhood (14, 15).

Several meta-analyses assessing DNA methylation of peripheral blood leukocytes of children with and without asthma have identified differentially methylated regions proximal, or within, genes specifically transcribed in eosinophils (16, 17). These findings are consistent with observations of enhanced Th2 responses in children with asthma (18). However, when assessing cord blood mononuclear cell (CBMC) samples, differential methylation did not extend to these and other classically Th2 associated loci (17). Additionally, randomized controlled trials using immunomodulatory products (e.g. Vitamin D, probiotics) in utero have failed to demonstrate benefit in prevention of asthma (19, 20). Thus, a greater understanding of early-life immunologic differences that are associated with predisposition to asthma may facilitate a more detailed risk stratification, and ultimately, identification of potential therapeutic targets.

Our focus was to determine biologic processes—extending to both transcriptional and serologic levels—associated with pediatric asthma risk that are detectable at birth. We hypothesized that transcriptional changes in CBMCs associated with multiple epidemiologic risk factors would be mediators of pediatric asthma risk. CBMCs have been previously studied with regards to cytokine production, DNA methylation, and outcomes (7, 15, 17, 18, 23), making them ideal candidates for investigation. Here, we identify a novel association between epidemiologic risk, neutrophil specific granules, and pediatric pulmonary outcomes.

## Results

### Approach to Identify Immunologic Differences Associated with Risk for Pediatric Asthma

We developed an analytic approach to identify genes whose expression in CBMCs are associated with newborns with higher or lower risk for asthma. We conducted a meta-analysis to increase power and generalizability (**Figure 1A, B**). In our approach, we queried NCBI’s Gene Expression Omnibus for CBMC microarray datasets that included metadata regarding the demographics and birth characteristics that are risk factors for pediatric asthma. We identified 354 datasets from our original search, of which 17 datasets contained relevant metadata. Of the 17 studies the most common maternal and neonatal characteristics reported were newborn sex, gestational age at birth, birthweight and maternal pre-pregnancy BMI (PP BMI) at 69.96%, 51.59%, 30.27%, and 19.32% of samples, respectively. Metadata regarding maternal smoking and mode of delivery (i.e., vaginal vs. cesarean section) was reported for 3 datasets; however, 2 of these datasets originated from the same laboratory group and contributed the majority of samples reporting these factors. None of the identified datasets reported metadata about maternal atopy or child’s ethnicity. Therefore, we chose to focus the analysis on gene expression associated with newborn sex, gestational age at birth, newborn weight and maternal PP BMI. 6 datasets were excluded due to homogenous metadata (e.g., all female). Datasets and the corresponding analysis are reported; a total of 605 unique transcriptomes were included (**Table 1**) (21-31).

**Table 1.**
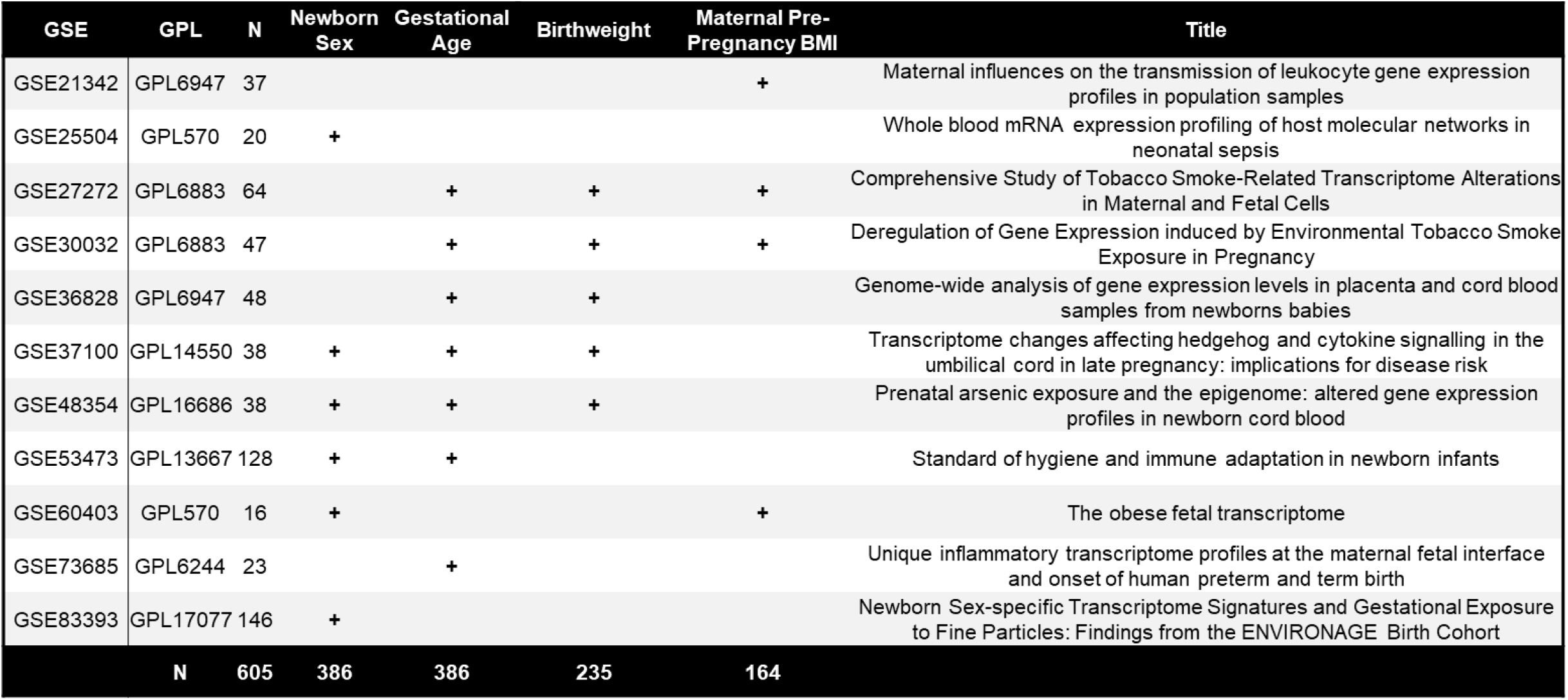
GSE Data Sets Used for Meta-Analysis.

| GSE | GPL | N | Newborn Sex | Gestational Age | Birthweight | Maternal Pre-Pregnancy BMI | Title |
| --- | --- | --- | --- | --- | --- | --- | --- |
| GSE21342 | GPL6947 | 37 |  |  |  | + | Maternal influences on the transmission of leukocyte gene expression profiles in population samples |
| GSE25504 | GPL570 | 20 | + |  |  |  | Whole blood mRNA expression profiling of host molecular networks in neonatal sepsis |
| GSE27272 | GPL6883 | 64 |  | + | + | + | Comprehensive Study of Tobacco Smoke-Related Transcriptome Alterations in Maternal and Fetal Cells |
| GSE30032 | GPL6883 | 47 |  | + | + | + | Deregulation of Gene Expression induced by Environmental Tobacco Smoke Exposure in Pregnancy |
| GSE36828 | GPL6947 | 48 |  | + | + |  | Genome-wide analysis of gene expression levels in placenta and cord blood samples from newborns babies |
| GSE37100 | GPL14550 | 38 | + | + | + |  | Transcriptome changes affecting hedgehog and cytokine signalling in the umbilical cord in late pregnancy: implications for disease risk |
| GSE48354 | GPL16686 | 38 | + | + | + |  | Prenatal arsenic exposure and the epigenome: altered gene expression profiles in newborn cord blood |
| GSE53473 | GPL13667 | 128 | + | + |  |  | Standard of hygiene and immune adaptation in newborn infants |
| GSE60403 | GPL570 | 16 | + |  |  | + | The obese fetal transcriptome |
| GSE73685 | GPL6244 | 23 |  | + |  |  | Unique inflammatory transcriptome profiles at the maternal fetal interface and onset of human preterm and term birth |
| GSE83393 | GPL17077 | 146 | + |  |  |  | Newborn Sex-specific Transcriptome Signatures and Gestational Exposure to Fine Particles: Findings from the ENVIRONAGE Birth Cohort |
| N |  | 605 | 386 | 386 | 235 | 164 |  |

**Figure 1.**
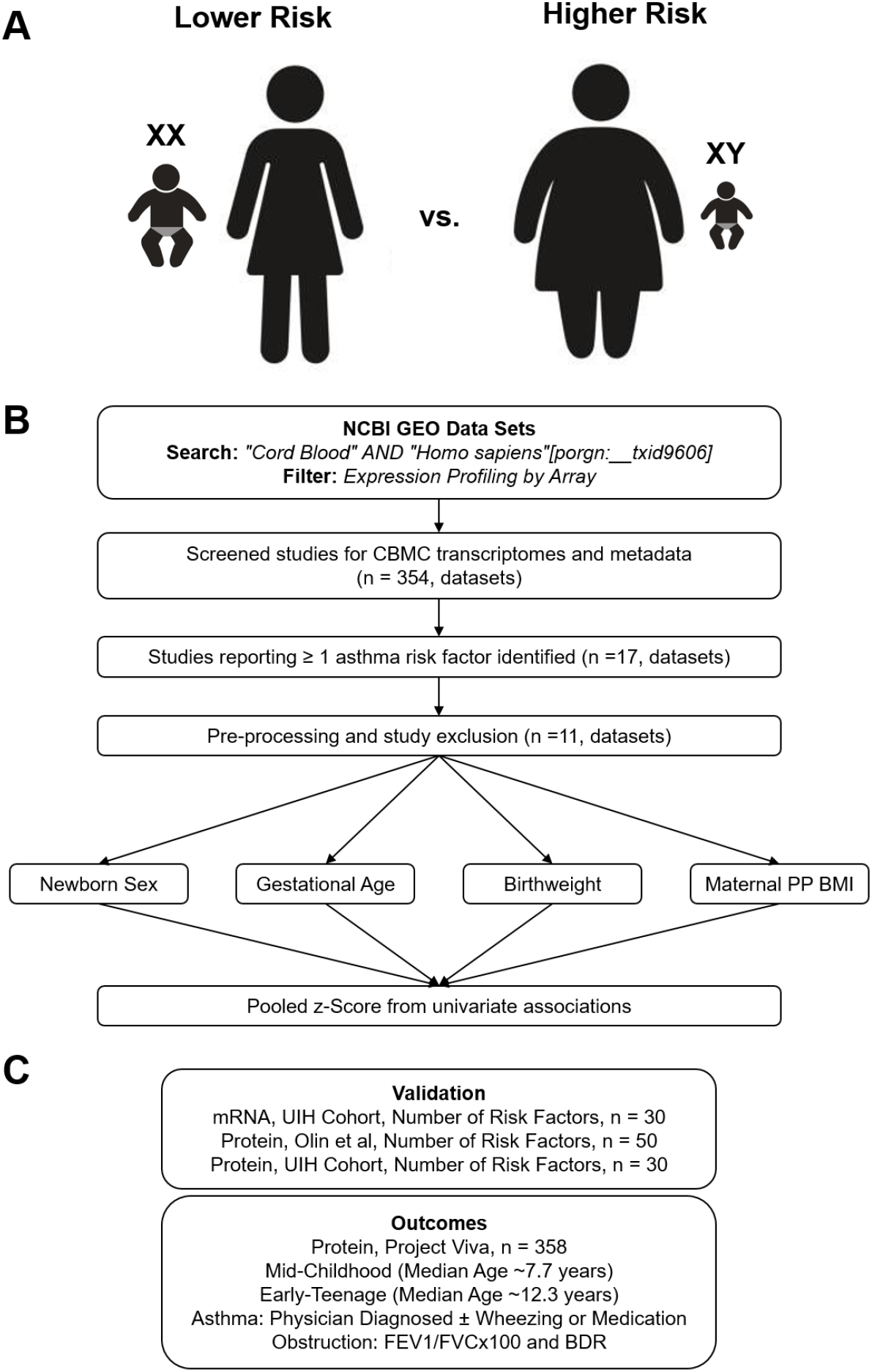
Overview of analytic approach used to identify biological risk for pediatric asthma. **(A)** Previously described perinatal risk factors for development of pediatric asthma: preterm birth, low birthweight, male, and maternal obesity. **(B)** Flow diagram of search, inclusion, exclusion, and univariate testing for transcriptomic analysis. **(C)** Cohorts, types of biosamples, and outcomes used for validation.

To validate the findings of the meta-analysis, we assessed three independent cohorts to confirm which genes are associated with asthma risk (**Figure 1C**). Further details regarding validation and outcomes follow in later result sections. In brief, the goal of validation was to assess gene expression in an independent cohort (UIH Cohort) where all subjects had complete metadata regarding newborn sex, gestational age, birthweight and PP BMI. We sought to further understand whether transcriptomic differences in CBMCs corresponded to differences at the protein level (all three cohorts) or cell population level (Olin et al (32)). Finally, we assessed 2 identified proteins in another independent cohort (Project Viva Cohort) to test for association with pediatric asthma and pulmonary function outcomes at 2 follow-up time points.

### Meta-Analysis of CBMC Transcription Associated with Individual Perinatal Risk Factors

Univariate random effects models were generated to assess transcriptional changes in CBMCs with regards to newborn sex, gestational age, birthweight, and maternal PP BMI (**Figure 2A**). Differential expression (FDR < 1%) was observed in 122, 34, 4, and 12 genes when comparing fetal sex, gestational age, birthweight, and maternal pre-pregnancy BMI, respectively (**Supplemental Data 1-4**). When evaluating sex and gestational age, several expected genes were identified to have large transcriptional changes. With regards to sex-associated transcriptional changes, although there was no X or Y chromosome-wide gene enrichment, several genes located on X (*KDM5C, SMC1A, TXLNG*, and *KDM6A*) and Y (*KDM5D* and *EIF1AY*) chromosomes exhibited the largest effect sizes and most significant differences. In addition to expected sex-associated transcriptional changes, *HBE1*, a hemoglobin subunit associated with fetal erythropoiesis, was significantly associated with pre-term gestational ages. To further validate our gestational age findings, we compared our univariate analysis with previously published results of differentially methylated regions associated with gestational age at birth estimated by last menstrual period (33) (**Figure S1**). A significant association was observed between differentially methylated genes and the effect size in our gestational age analysis, such that genes whose methylation increased with gestational age as reported by Bohlin et al. (33) showed on average decreased expression with gestational age in our meta-analysis.

**Figure 2.**
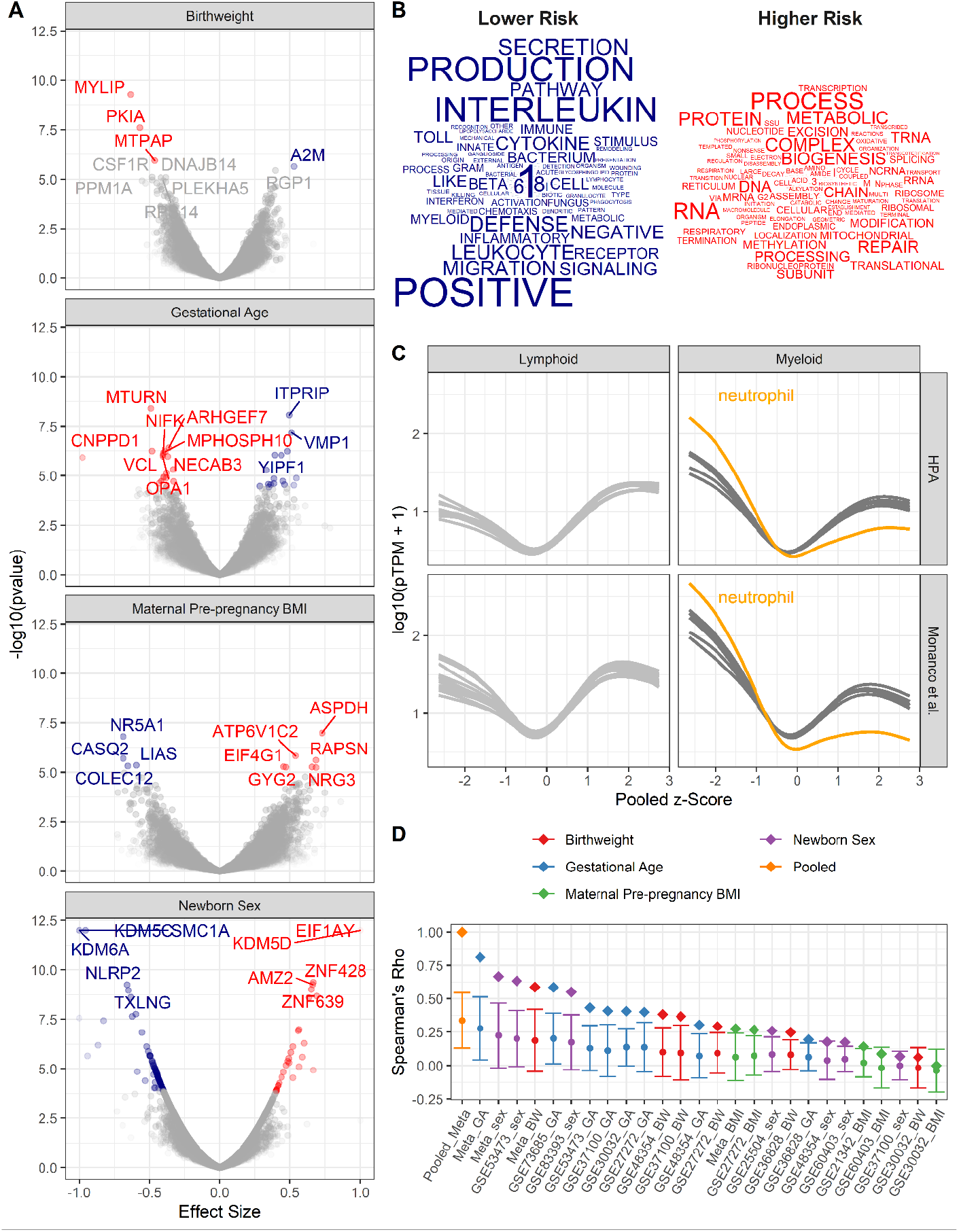
Pooled meta-analysis z-scores identify gene expression signatures related to asthma risk. Significant (FDR < 1%) genes and gene sets are colored by their association with either higher (red) or lower (blue) risk. **(A)** Volcano plots of gene expression for univariate analyses. Top 10 most significant genes labeled. **(B)** Word clouds of GO terms significantly enriched (FDR<1%) using the pooled z-score as pre-ranked list for GSEA. **(C)** Protein coding transcripts per million reads (pTPM) in peripheral blood cells (Human Protein Atlas and Monaco et al., (34, 35)) relative to pooled z-score. Each line represents one cell type; neutrophils highlighted in orange. **(D)** Spearman’s correlation between pooled z-statistic and individual analyses (diamonds). Average Spearman’s correlations between individual analyses and combination of all other analyses (circle), SD indicated by error bars.

### Pooled Meta-Analysis Gene Expression Signature

To identify the biological processes that are enriched by genes with transcriptional changes associated with higher or lower risk of asthma, the z-scores from each univariate meta-analysis were averaged, such that negative z-statistics were associated with lower risk (female, older gestational ages, higher birthweights, and lower maternal PP BMI) and positive z-statistics were associated with increased risk. Thus, the pooled z-score indicates the average probability that a gene’s expression is associated with either increased or decreased risk of asthma based on an individual’s demographics and birth characteristics. The averaged z-statistic was used as a pre-ranked list for gene set enrichment analysis. GO terms were assessed for enrichment; 18 and 19 GO terms were significantly enriched (FDR < 1%) with regards to low and high risk profiles, respectively (**Figure 2B**). Genes associated with lower risk exhibited increased representation in GO terms involving innate immune signaling and defense, whereas high risk genes were enriched in pathways involving translation and RNA metabolic processes.

Gene expression studies from pooled cellular populations (e.g. CBMCs, PBMCs, and tissues) can be influenced by the cellular composition. To determine if specific cell population enrichment was associated with the pooled z-score, the Human Protein Atlas (HPA, Monaco et al.) (34, 35) was utilized to assess the abundance of transcripts in peripheral blood leukocyte RNA transcriptomes in relationship to the pooled z-score (**Figure S2**). We observed a generalized increase in expression of low risk genes in myeloid cells and high risk genes in lymphoid cells. This pattern of expression was most pronounced in neutrophils (**Figure 2C**). These results suggest that lower risk individuals have increased populations of myeloid cells in their CBMCs.

A potential confounder in pooling results is the potential over-representation of any one analysis. To assess bias in the pooled z-score, two analyses were performed (**Figure 2D**). In the first assessment, z-scores from each individual analysis correlated with the pooled z-score. The highest correlations with the pooled z-score were the z-scores from the meta-analyses assessing gestational age at birth, newborn sex, and birthweight. Individual dataset z-scores for each dataset demonstrated a similar trend. In the second assessment, z-scores from each individual analysis were correlated with the combination of all other z-scores. Again, the pooled z-score had the highest average correlation followed by the meta-analyses. Together, this demonstrates that the pooled z-score does, indeed, amalgamate information across all of the analyses, with the most influence arising from gestational age at birth, newborn sex, and birthweight.

### Specific Granule Gene Expression Association with Multiple Pediatric Asthma Risk Factors

To confirm gene expression changes associated with asthma risk stratification, pooled z-scores from the meta-analysis were compared with the UIH cohort, a cohort of individuals in which newborn sex, maternal PP BMI, gestational age at birth, and birthweight were known (**Supplemental Data 5**). UIH cohort z-scores were calculated from mRNAseq of CBMCs, where gene expression was modeled as a function of number of risk factors (**Supplemental Data 6**). We developed a method to validate the congruence between the UIH cohort and the pooled meta-analysis, which we termed the replication score (RS). This RS is the product of the pooled z-score from the meta-analysis and the UIH z-score (see methods). We assessed the relationship between RS cutoff, p-values, and number of genes (**Figure S2**). Genes with a replication score greater than 3 were identified as being sufficiently congruent. 51 genes, 0.4% of all genes tested had a RS greater than 3 (**Figure 3A**). These identified genes corresponded well with the results of pooled z-score for gestational age at birth, newborn sex and birthweight, but showed limited correlation to maternal PP BMI. They had median p-values of 0.02, 0.01, 0.11, and 0.58 in the gestational age at birth, newborn sex, birthweight, and maternal PP BMI meta-analyses, and median p-value of 0.02 in the UIH cohort.

**Figure 3.**
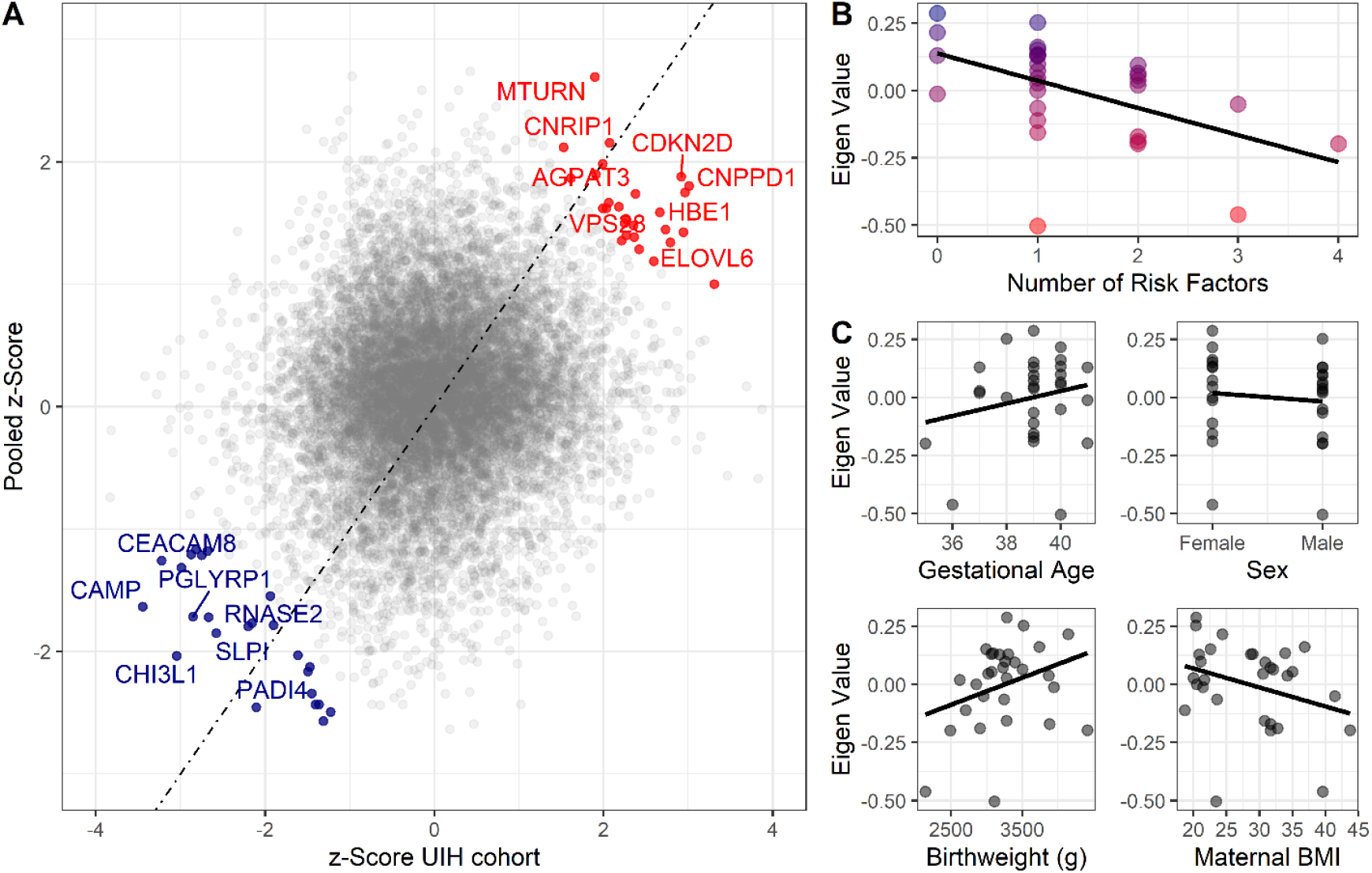
Validation cohort identifies gene signature associated with pediatric asthma risk factors. Color labeling indicating association with either higher (red) or lower (blue) risk of pediatric asthma development **(A)** Dot-plot demonstrating validation between meta-analysis Pooled z-score and UIH cohort mRNAseq z-score. Colored and labeled dots indicate those with non-parametric replication score greater than 3 and 4, respectively. **(B-C)** Association between number of risk factors or individual risk factors and eigenvalue of gene signature (validation score >3), UIH Cohort.

Replicating genes were enriched for processes involved in vesicle biology (**Figure S3**). Specifically, replicating genes associated with low risk were enriched for genes that are components of granulocyte specific granules (*MS4A3, CEACAM8, OLR1, CAMP, LTF, CHI3L1, SLPI, PGLYRP1*). With regards to genes associated with higher risk, genes involved in vesicle sorting/production (*VPS28, VTI1B, FIS1*) as well as several genes involved in vesicle membrane biology (*AGPAT3, ELOVL6, TM7SF2*) were identified.

To test whether replicating genes were associated with the number of risk factors, these 51 genes were assessed using principal component analysis. Using the first Eigenvector (explaining 41.3% of variance in replicating genes), a significant association (R [95%CI] = -0.51[-0.73, -0.18], p-value < 0.01) was observed between UIH cohort Eigenvalues and number of epidemiologic risk factors (**Figure 3B**). Positive eigenvalues represent increased expression of low risk genes and negative eigenvalues represent increased expression of high risk genes. Although the associations with individual risk factors were in the expected directions, they were not significant (**Figure 3C**), suggesting that the additive effect is greater than the individual.

### Cellular and Protein Abundance in Relation to Pediatric Asthma Risk Factors

To further determine if these changes are due to differences in cellular populations, we analyzed mass cytometry data published by Olin et al. for abundance of 21 different cell types in relationship to number of epidemiologic risk factors for pediatric asthma (32). Cord blood neutrophil abundance was inversely associated (R [95%CI] = -0.57 [-0.73, -0.34], Bonferroni p-adj <0.001) with the number of risk factors (**Figure 4A**). Other myeloid cell types, CD14+ monocytes and myeloid-derived dendritic cells, also had negative correlations but were weaker and not significant after multiple testing correction (**data not shown**).

**Figure 4.**
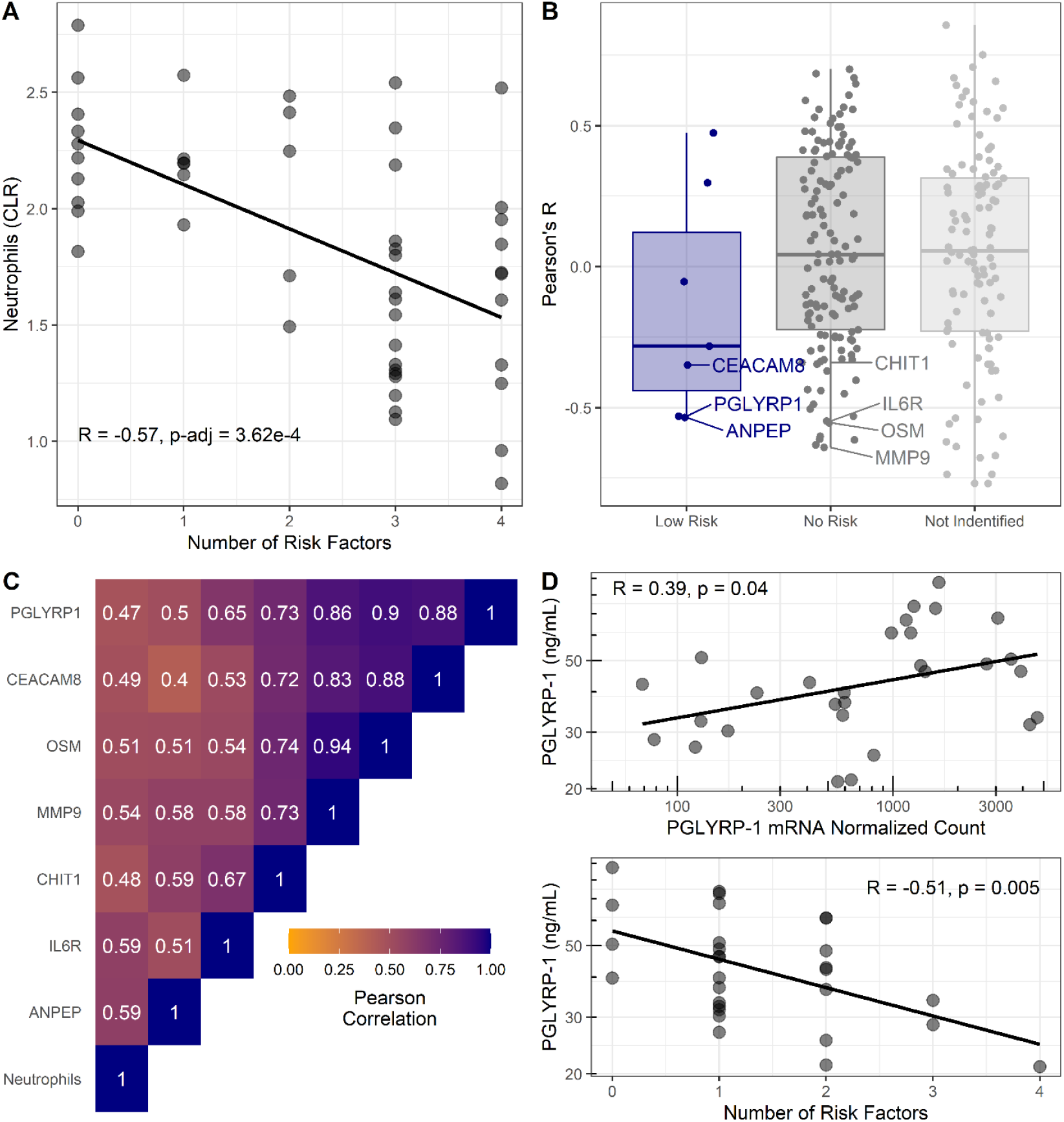
Cellular and proteomic differences associated with pediatric asthma risk factors. **(A-C)** Re-analysis of publicly available data from Olin et al. (32). **(A)** Percentage of neutrophils in cord blood (transformed using centered log-ratios, CLR) correlated with number of risk factors. Pearson’s Correlation (R) and Bonferroni adjusted p-value reported. **(B)** Pearson’s correlation coefficients (R) for plasma-protein concentration and number of risk factors distributed based on risk association of proteins as per Figure 3. Corresponding mRNA from CBMCs were identified for low risk associated proteins (blue) and no risk associated proteins (dark grey). Most significant negative protein correlations with neutrophil-enriched mRNA (Human Protein Atlas, (34)) are notated. Proteins identified in previous analysis without corresponding mRNA shown light grey. **(C)** Heatmap of Pearson’s correlations between neutrophils and neutrophil-derived proteins identified in (B). **(D)** Association between PGLYRP-1 umbilical cord serum concentration, PGLYRP-1 CBMC mRNA, and number of risk factors in UIH cohort.

Extending these findings from gene expression to protein abundance, reported umbilical cord plasma protein abundance data was correlated with number of risk factors in a secondary analysis (32). Serum proteins from genes that replicated with low risk had on average negative correlations with number of risk factors (**Figure 4B**). No proteins from the high risk genes were tested in plasma, due to their intracellular localization. Notably, proteins (CEACAM8, PGLYRP-1, CHIT1, sIL6Rα, MMP-9, and OSM) predicted to be enriched in neutrophils by the HPA (34) had strong correlations with both neutrophil abundance and number of risk factors (**Figure 4B-C**).

We hypothesized that the serum concentration of proteins identified as low risk in our transcriptomic analysis would correlate with mRNA abundance in CBMCs, whereas those not associated with risk would not correlate with mRNA in CBMCs. To test this hypothesis, we used the UIH cohort to correlate mRNA abundance with serum protein concentration of PGLYRP-1 (low risk) and sIL6Rα (no risk). We observed a significant (R [95%CI] = 0.39 [0.03, 0.66], p<0.05) association between PGLYRP-1 protein concentration and mRNA (**Figure 4D**). Consistent with transcriptomic results, PGLYRP-1 cord blood serum concentration was inversely associated with number of risk factors (R [95%CI] = -0.51 [-0.74, -0.17], p<0.01). sIL6Rα was neither associated with its mRNA in CBMCs (R [95%CI] = 0.37 [-0.22, 0.77], p=0.21) nor the number of risk factors (R [95%CI] = 0.20 [-0.39, 0.67], p=0.50).

### Demographic Associations with Serum Neutrophil Proteins in UIH and Project Viva Cohorts

We tested the association between PGLYRP-1 and sIL6Rα, individual risk factors and demographics in the UIH and Project Viva cohorts. Cord blood serum was available in a subset of individuals (n = 358) from Project Viva (**Supplemental Data 7**). There was no significant difference (p>0.05, Wilcoxon rank sum test) in PGLYRP-1 or sIL6Rα between UIH and Project Viva cohorts (**Figure S4**). Consistent with our previous observations, PGLYRP-1 and sIL6Rα were positively correlated in both UIH (R [95%CI] = 0.21 [-0.16, 0.54]) and Project Viva (R [95%CI] = 0.19 [0.09, 0.29]) cohorts. Although there were no significant associations between PGLYRP-1 or sIL6Rα with any single demographic in all three data sets, there was a small positive association between PGLYRP-1 and gestational age at birth (**Table 2**). Collectively, these results suggest that increased abundance of mRNA from genes localizing to neutrophil specific granules are associated with the number of risk factors for pediatric asthma. These changes in mRNA are reflected in the abundance of these specific granule proteins in serum and plasma.

**Table 2.**
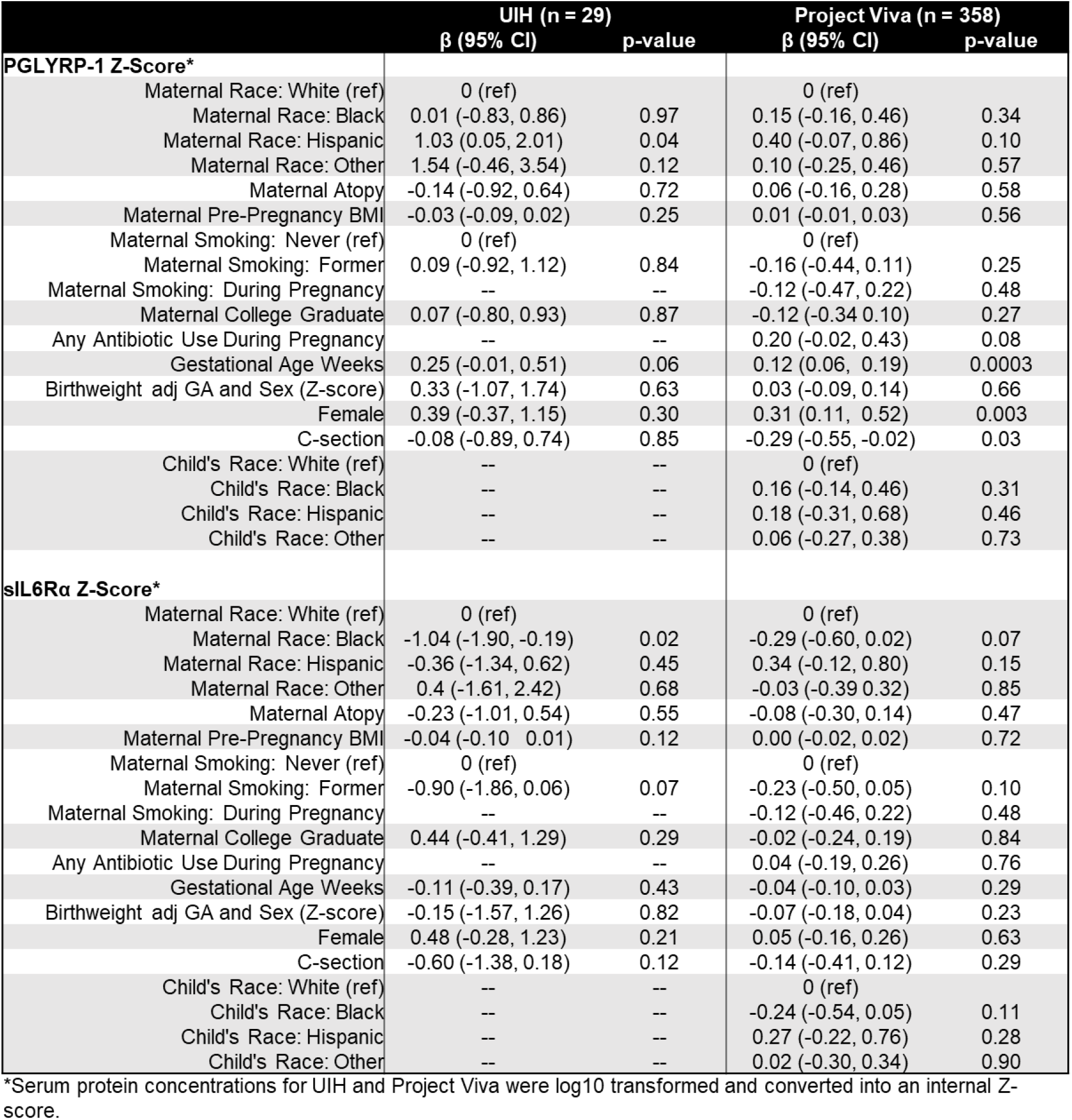
Univariate Associations between Cord Blood Serum Proteins and Covariates.

### Serum Neutrophil Proteins Associations with Pediatric Pulmonary Outcomes

In context of our previous results, we hypothesize that specific granule protein abundance in serum is associated with risk of pediatric asthma and this process is independent of neutrophil abundance. To evaluate this hypothesis, we measured PGLYRP-1 (present in neutrophil specific granule and correlates with its mRNA in CBMCs) and sIL6Rα (derived from neutrophils but not present in specific granules and does not correlate with its mRNA in CBMCs) in umbilical cord blood serum. At two follow up time-points, asthma outcomes and expiratory flow volumes were modeled as a function of PGLYRP-1 and sIL6Rα in a subset of individuals in Project Viva (**Figure S5**). The demographics of the subset of individuals from Project Viva from which umbilical cord serum was available had a similar demographic profile as the full cohort. One notable difference was a sizeable decreased response rate for asthma outcomes at the early-teenage follow-up compared to the full cohort (32% subset vs. 47% full cohort).

PGLYRP-1 and sIL6Rα were modeled as predictors for current asthma at mid-childhood (median age ∼7.7 years old) and early-teen (median age ∼12.8 years old) follow-ups (**Table 3**). Four regression models were used to estimate the association between asthma outcomes: univariate, adjustment for child’s birth characteristics and demographics, adjustment for mother’s demographics, and adjustment for birth characteristics and all demographics (reported in manuscript). The abundance of PGLYRP-1 was significantly associated with current asthma at mid-childhood (adjusted OR [95% CI]: 0.50 [0.31, 0.77] per 1SD increase, p-value = 0.003) (**Figure 5A**). There were no significant associations between current asthma and PGLYRP-1 at the early-teen follow-up, however the confidence interval at this time-point was much wider, likely secondary to the smaller sample size. There were no significant association between sIL6Rα with any asthma outcome at either time-point.

**Table 3.** Association between Cord Blood Serum Proteins and Asthma Outcomes.

|  | Mid-Childhood |  | Early-Teen |  |
| --- | --- | --- | --- | --- |
|  | Current Asthma<br>OR (95% CI) | Ever Asthma<br>OR (95% CI) | Current Asthma<br>OR (95% CI) | Ever Asthma<br>OR (95% CI) |
| **PGLYRP-1**<br>**Z-Score\*** |  |  |  |  |
| Univariate | 0.52 (0.35, 0.75) | 0.52 (0.36, 0.74) | 0.65 (0.39, 1.10) | 0.64 (0.45, 0.89) |
| Model 1<sup>†</sup> | 0.57 (0.37, 0.85) | 0.54 (0.36, 0.79) | 0.86 (0.48, 1.54) | 0.72 (0.50, 1.03) |
| Model 2<sup>#</sup> | 0.57 (0.26, 0.65) | 0.41 (0.26, 0.61) | 0.62 (0.35, 1.09) | 0.61 (0.42, 0.87) |
| Model 3<sup>##</sup> | 0.50 (0.31, 0.77) | 0.48 (0.31, 0.77) | 0.88 (0.45, 1.72) | 0.74 (0.51, 1.07) |
| **sIL6Rα**<br>**Z-Score\*** |  |  |  |  |
| Univariate | 0.87 (0.61, 1.23) | 0.93 (0.67, 1.29) | 0.75 (0.42, 1.28) | 0.93 (0.66, 1.29) |
| Model 1<sup>†</sup> | 0.83 (0.56, 1.23) | 0.89 (0.63, 1.30) | 0.68 (0.37, 1.21) | 0.90 (0.63, 1.27) |
| Model 2<sup>#</sup> | 0.83 (0.57, 1.24) | 0.90 (0.63, 1.28) | 0.67 (0.33, 1.27) | 0.93 (0.65, 1.31) |
| Model 3<sup>##</sup> | 0.83 (0.55, 1.25) | 0.90 (0.62, 1.30) | 0.60 (0.27, 1.19) | 0.88 (0.61, 1.27) |

\* Serum protein concentrations were log10 transformed and converted into an internal Z-score
† (Child's Demographics and Birth Characteristics): adjusted for gestational age, birthweight adjusted for gestational age, mode of delivery, child's sex, child's race/ethnicity

**Figure 5.**
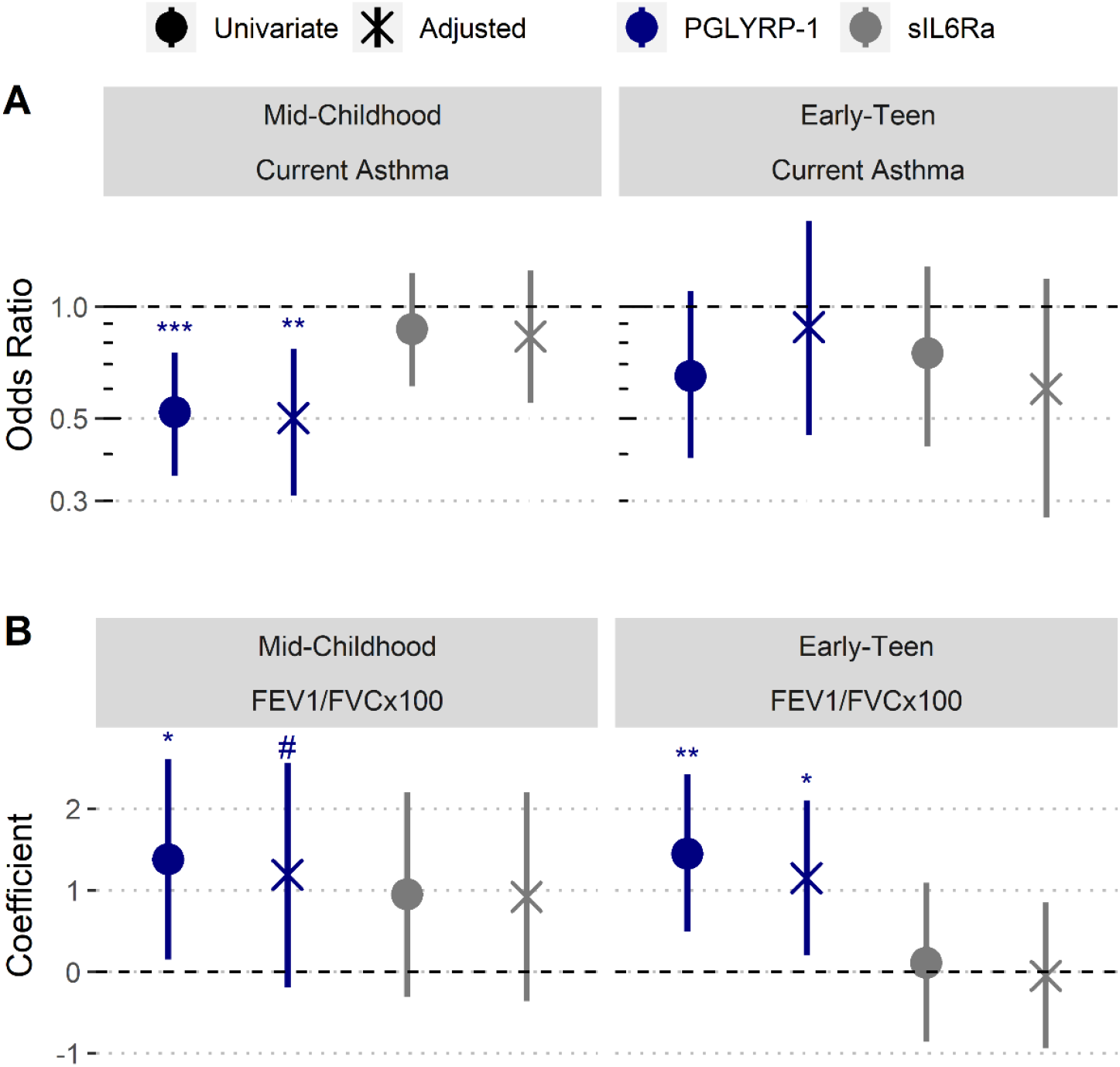
Increased umbilical cord blood serum PGLYRP-1 is associated with increased FEV1/FVC and reduced odds of pediatric asthma. Samples and data derived from a subset of Project Viva (n= 358). Odds ratio and coefficient estimates are based on one standard deviation increase in serum proteins (PGLYRP-1, sIL6Rα). Error bars indicate 95% CI. Adjusted model co-variates: gestational age, birthweight adjusted for gestational age and sex, mode of delivery, child’s sex, child’s race/ethnicity, maternal pre-pregnancy BMI, maternal level of education, maternal atopy, antibiotic exposure during pregnancy, and early life smoke exposure. **(A)** PGLYRP-1 and sIL6Rα concentrations in umbilical cord blood serum associations with current asthma at mid-childhood and early-teenage time points (determined by questionnaire responses). **(B)** PGLYRP-1 and sIL6Rα concentration in umbilical cord blood serum associations with FEV1/FVC×100 at mid-childhood and early-teenage follow-ups. **\*\*\*** p< 0.001, **\*\*** p<0.01, **\*** p<0.05, **#** p<0.1

We also performed analyses to estimate the relationship of cord blood PGLYRP-1 and sIL6Rα with FEV_1_/FVC×100 ratio and bronchodilator response (BDR) at mid-childhood and early-teen follow-up time points (**Table 4**). There was no significant association between sIL6Rα and FEV_1_/FVC×100 ratio at either time-point. PGLYRP-1 and sIL6Rα were not associated with BDR at either time-point. However, there was trend towards an association between PGLYRP-1 concentration and FEV_1_/FVC×100 ratio at mid-childhood (adjusted β [95% CI]: 1.18 [-0.18, 2.56] per 1SD increase, p-value = 0.09) and significant association at the early-teen follow up (adjusted β [95% CI]: 1.15 [0.20, 2.10] per 1SD increase, p-value = 0.02) (**Figure 5B**).

**Table 4.** Association between Cord Blood Serum Proteins and Pulmonary Function Tests.

|  |  | Mid-Childhood |  | Early-Teen |  |
| --- | --- | --- | --- | --- | --- |
|  |  | FEV1/FVCx100 | BDR | FEV1/FVCx100 | BDR |
| | | $\beta$ (95% CI) | $\beta$ (95% CI) | $\beta$ (95% CI) | $\beta$ (95% CI) |
| **PGLYRP-1 Z-Score\*** |  |  |  |  |  |
|  | Univariate | 1.38 (0.15, 2.61) | 0.10 (-1.83, 2.03) | 1.45 (0.49, 2.42) | -0.68 (-1.54, 0.17) |
|  | Model 1<sup>†</sup> | 1.12 (-0.18, 2.41) | 0.53 (-1.44, 2.51) | 1.05 (0.11, 1.98) | -0.49 (-1.40, 0.41) |
|  | Model 2<sup>#</sup> | 1.38 (0.08, 2.69) | 0.61 (-1.40, 2.63) | 1.53 (0.54, 2.53) | -0.58 (-1.49, 0.34) |
|  | Model 3<sup>##</sup> | 1.19 (-0.19, 2.56) | 1.00 (-1.04, 3.04) | 1.15 (0.20, 2.10) | -0.35 (-1.29, 0.59) |
| **sIL6Rα Z-Score\*** |  |  |  |  |  |
|  | Univariate | 0.95 (-0.31, 2.20) | -0.38 (-2.45, 1.69) | 0.11 (-0.86, 1.09) | 0.65 (-0.24, 1.54) |
|  | Model 1<sup>†</sup> | 0.96 (-0.30, 2.21) | -0.37 (-2.41, 1.67) | 0.02 (-0.87, 0.92) | 0.71 (-0.19, 1.61) |
|  | Model 2<sup>#</sup> | 0.88 (-0.40, 2.16) | -0.62 (-2.62, 1.39) | -0.02 (-1.00, 0.97) | 0.75 (-0.17, 1.66) |
|  | Model 3<sup>##</sup> | 0.92 (-0.36, 2.20) | -0.11 (-0.96, 0.70) | -0.05 (-0.94, 0.85) | 0.79 (-0.12, 1.69) |

\* Serum protein concentrations were log10 transformed and converted into an internal Z-score
† (Child's Demographics and Birth Characteristics): adjusted for gestational age, birthweight adjusted for gestational age, mode of delivery, child's sex, child's race/ethnicity

To further our understanding of the relationship between PGLYRP-1 and outcomes, we performed two secondary analyses of models adjusted for all demographics and birth characteristics. First, we assessed the total variance explained by the regression model adjusting for both birth characteristics and demographics, and the variance explained by each of the individual predictors in the model. Assessing current asthma risk at mid-childhood, the regression model explained approximately 18% of the variance, and PGLYRP-1 was the most important predictor (**Figure S6**). Assessing current FEV1/FVC×100 at early-teen time-point, the model explained approximately 26% of variance, and PGLYRP-1 was the second most important predictor (**Figure S6**). Second, to identify covariates that modify the effect of PGLYRP-1, we performed subset analyses (**Figure S7**). Small for gestational age and children identified by their mothers as “Other” race/ethnicity displayed significantly different associations between PGLYRP-1 and mid-childhood asthma. Small for gestational age and children with obese mothers displayed significantly different associations between PGLYRP-1 and FEV1/FVC.

These findings implicate a role for PGLYRP-1 and other specific granule proteins as predictors of pediatric asthma risk and pulmonary function. This is in contrast to sIL6Rα, which is not localized to specific granules, its protein abundance is not regulated by transcription, and is not associated with any pulmonary outcomes.

## Discussion

Our study has identified a novel association between epidemiologic risk, neutrophil specific granules, and pediatric pulmonary outcomes. By pooling the meta-analysis results, we established an association between multiple risk factors and the expression of genes involved in innate immunity and nucleic acid metabolism. We hypothesized that this gene signature represents increased myelopoiesis *in utero* and correlates with perinatal risk for pediatric asthma. During the process of myeloid cell differentiation, production of proteins responsible for defense against microbes and pro-inflammatory signaling (e.g., IL-1β) are amplified, while translational activity and nucleolar size wane (36, 37). In our analysis, lower risk genes had higher expression in myeloid cells, most notably neutrophils. The low risk genes were enriched for those that are implicated in defense responses towards bacteria and fungi. Additionally, this gene signature was strongly correlated with gestational age at birth, newborn sex, and birthweight (weaker association with maternal PP BMI). Our findings parallel previous literature, which has demonstrated that preterm birth, male sex, and low birthweight are associated reduced abundance of neutrophils and monocytes (10).

Our replication of the pooled meta-analysis with the UIH cohort pointed towards genes located and involved in the biology of neutrophil specific (secondary) granules. In particular, lower risk individuals had higher expression of genes whose protein products are luminal (*PGLRYP1, LTF, PTX3, CHI3L1, CAMP, SLPI*) and membrane (*CEACAM8, MS4A3, OLR1*) components of specific granules (38). It is known that gestational age at birth, sex, mode of delivery, and birthweight may influence not only the abundance of neutrophils, but also their functionality (e.g. phagocytosis, cytokine production, respiratory burst) (39). We demonstrate that the additive effect of multiple risk factors is associated reduction of transcription and protein products of specific granules in umbilical cord blood serum. Notably, our reanalysis of mass cytometry and proteomic data demonstrated a correlation between PGLYRP-1 in serum and neutrophil abundance (32). Previous literature has shown that deficiency of SLPI leads to impairment of neutrophil development and abundance (40). Further, individuals with specific granule deficiency syndrome have abnormal neutrophil morphology, increased susceptibility to infections, and increased risk of acute myeloid leukemia (41). Thus, it is likely that neutrophil differentiation and survival is partially dependent on secondary granule generation.

To further investigate how these findings are related to pediatric asthma, we chose to compare PGLYRP-1, sIL6Rα, and their associations with asthma and lung function outcomes. PGLYRP-1 and sIL6Rα were chosen because both were correlated with neutrophil abundance, yet they are derived from different processes. Variation in abundance of PGLYRP-1 in serum is due to changes in neutrophil degranulation and transcription of *PGLYRP1*, whereas sIL6Rα is derived from receptor shedding and differential splicing of its mRNAs (42, 43). Thus, if perinatal neutrophil abundance is associated with pediatric asthma both PGLYRP-1 and sIL6Rα should demonstrate associations with these outcomes. In contrast, we observed a significant relationship between mid-childhood asthma and PGLYRP-1 not sIL6Rα, indicating that serum abundance of specific granule contents is associated with risk and not the abundance of cord blood neutrophils.

Whether PGLYRP-1 has a causal role in asthma pathogenesis or is merely a biomarker for risk remains to be determined. PGLYRP-1 has antimicrobial function, although the concentration we observed in cord blood is below those reported for *in vitro* studies (44-46). PGLYRP-1 functions synergistically *in vitro* with other antimicrobials (e.g. lysozyme), so potential benefit as an antimicrobial cannot be ruled out (45, 46). Interestingly, mammalian PGLYRP-1 does not hydrolyze peptidoglycan and its orthologues appear divergent from ancestral PGLYRPs which contain enzymatic activity, thus it may have roles other than that of an antimicrobial (47). It is also possible that other individual specific granule proteins or their cumulative effects are associated with pediatric asthma. Further studies will be required to determine a mechanistic role for either PGLYRP-1 or other specific granule proteins.

There are several murine studies linking PGLYRP-1 to increased airway resistance and perturbed immunity in response to house dust mite (48, 49). These murine studies have utilized adult mice deficient in PGLYRP-1 via germ-line deletion. One study has demonstrated that bone marrow reconstitution with WT bone marrow prior allergen sensitization abrogates this effect (49), suggesting PGLYRP-1 presence in adult mice at the time of sensitization is responsible for increased airway resistance and perturbed immunity. This is not surprising as PGLYRP-1 is reported to be a pro-inflammatory TREM-1 ligand and thus may propagate inflammation (43). Furthering this point, there are reported associations between increased serum PGLYRP-1 and systemic inflammatory conditions in adulthood (e.g. rheumatoid arthritis, cardiovascular disease) (50, 51). Notably, PGLYRP-1 and many of the other specific granule proteins are dramatically reduced in serum concentration one week postnatal (32). Murine studies using conditional knockouts at birth would better assess how the natural variation of PGLYRP-1 effects the susceptibility to pediatric asthma.

Although our findings contribute to our understanding of the risk for pediatric asthma, there are limitations. At the early-teen follow up, we did not observe a significant association with current asthma. It is important to note that there was still an association between PGLYRP-1 and FEV1/FVC at this time-point. These observations could be due two possibilities. First, at early-teen follow-up, there was only 9% prevalence of current asthma in the Project Viva subset, whereas the full cohort had a 15% prevalence. This difference is likely due to higher non-response rates in the asthmatic sub-group as 24 of the 35 (68%) had missing responses, whereas only 46 of 171 (27%) of non-asthmatics had missing responses. This lead to a reduction in power at the early-teen follow-up time point, potentially leading to a false negative result. Second, PGLYRP-1 might be inversely associated with FEV1/FVC in adolescence secondary to early-life pulmonary dysfunction (e.g. mid-childhood asthma). Reduced expiratory pulmonary function later in adolescences and even into adulthood is associated with individuals who were diagnosed with asthma in childhood (52-55). We view these possibilities as equally probable as current pediatric asthma can also impair FEV1/FVC (56). Further investigation will be need to elucidate the role of PGLYRP-1 in adolescent asthma.

In conclusion, we have identified a neutrophil development gene signature that is associated with perinatal asthma risk. A soluble specific granule protein, PGLYRP-1, was strongly associated with odds of asthma in childhood and pulmonary function in childhood and adolescence. This suggests that perinatal granulopoiesis has a significant impact on the development of pediatric asthma and lung function.

## Methods

### Human Subjects Project Viva Cohort

The current study was approved by the University of Illinois at Chicago IRB (#2016–0326) and the IRB of Harvard Pilgrim Health Care. Volunteers were recruited from women attending their first prenatal visit at one of 8 practices of Atrius Harvard Vanguard Medical Associates. The exclusion criteria were multiple gestation, inability to answer questions in English, gestational age ≥ 22 weeks at recruitment, and plans to move away before delivery. The cohort profile was previously described by Oken and colleagues (57).

Current mid-childhood asthma was defined as a “yes” response to “Has a health professional ever told you that your child has asthma?” and “yes” to either “In the past 12 months, has child taken or been prescribed Albuterol, Cromolyn, Nedocromil, Montelukast, inhaled corticosteroids, or Prednisone” or “In the past 12 months, has your child ever had wheezing (or whistling in the chest)?”. We used as a comparison group those with no asthma diagnosis and no asthma medication use or wheezing in the past 12 months. We used the same definition for current asthma in early adolescence, except the time reference for asthma medication was “in the past month”. Ever asthma was defined as a “yes” response to “Has a health professional ever told you that your child has asthma?” within either the mid-childhood or early-teen follow up periods. We used as a comparison group those with no asthma diagnosis. Individuals with missing data were not used in regression models assessing current or ever asthma outcomes. Individuals with missing data are reported in demographics tables and displayed in figure as “Missing Data”.

Methods for obtaining spirometric measurements and bronchodilator response have been described previously (56). In brief, spirometry was performed with the EasyOne Spirometer (NDD Medical Technologies, Andover, Mass). Post-bronchodilator spirometric measures were obtained at least 15 minutes after administration of 2 puffs (90 μg per puff) of albuterol. Spirometric performance was required to meet American Thoracic Society criteria for acceptability and reproducibility, with each subject producing at least 3 acceptable spirograms, 2 of which must have been reproducible (57, 58).

### Human Subjects University of Illinois Hospital Cohort

The current study was approved by the University of Illinois at Chicago IRB (#2015–0353). Women seeking prenatal care at the University of Illinois at Chicago (UIC) Center for Women’s Health were recruited as volunteers in their third trimester (29–33 weeks of gestation) between 2014 and 2017. The cohort profile has been previously described by Koenig and colleagues (59). Inclusion criteria as follows: singleton pregnancy; naturally conceived pregnancy; 17–45 y of age; pre-pregnancy BMI ≥18.5; < 34 weeks of gestation; sufficient fluency in English to provide consent and complete the study; and ability to independently provide consent. Exclusion criteria as follows: live birth or another pregnancy (including ectopic and molar pregnancies) in the previous 12 months; preeclampsia; gestational diabetes mellitus or previously diagnosed type 1 or type 2 diabetes; autoimmune disorder; current or previous premature rupture of membranes or chorioamnionitis; previous spontaneous premature birth; current bacterial or viral infection; current steroid or anti-inflammatory treatment; history of bariatric surgery; malabsorptive condition (e.g., celiac disease); current hyperemesis; hematologic disorder (e.g., sickle cell anemia or trait, hemochromatosis); current tobacco use; alcohol consumption or illicit drug use; and current use of medications that decrease nutrient absorption (e.g., proton pump inhibitors). All women provided written informed consent.

### Umbilical Cord Blood Specimens

For Project Viva, procedures for obtaining umbilical cord blood serum have been describe previously (60). For the UIH cohort, umbilical cord blood was obtained by venipuncture shortly after time of delivery. Blood (approximately 5 mL per tube) was drawn into Red Top Serum Plus and Green Top Sodium Heparin 95 USP Units Blood Collection Tubes (BD Vacutainer). Red Tops were allowed to stand upright at room temperature for 30 min prior centrifugation at 1500x g for 10 minutes at room temperature. Supernatants (serum) were collected, aliquoted, and stored at - 80C until further processing. Heparinized blood obtained in Green Tops was diluted 1:1 in 1x Phosphate Buffered Saline, pH 7.4 (PBS) and overlaid onto Ficoll-Paque^™^ Plus (GE Healthcare) density gradients. Density gradients were centrifuged at 400x g for 30 minutes at room temperature without brake. Upper phase (diluted plasma) was drawn off, aliquoted, and stored at -80C. Buffy coats (CBMCs) were drawn off, washed twice with 10 mL of 1x PBS, aliquoted and stored in 500 mL of RNAlater (Qiagen). Viability and number of cells isolated was determined by diluting cellular suspensions 1:1 with Trypan Blue Solution 0.4% (w/v) in PBS (Corning) and counting live/dead cells >7 μM using a TC20^™^ Automated Cell Counter (Biorad). The viability of isolated cells was >90% for all samples. Time from delivery to storage was recorded for every sample.

### RNA Extraction and Sequencing

Total RNA was extracted from CBMCs using RNeasy kits (QIAGEN) following manufacturers protocol except for switching 70% ethanol for 100% ethanol. The quality and quantity of all the extracted RNA was analyzed with a RNA 6000 Nano Kit on the 2100 Bioanalyzer Instrument (Agilent) and ssRNA High Sensitivity Kit and Qubit (Invitrogen). RIN for all samples was > 8. RNA was constructed into barcoded libraries using the TruSeq Stranded mRNA Library Prep Kit (Illumina). The pooled libraries were sequenced for a paired-end 151 read length. The DNA libraries were sequenced on HiSeq X Ten platform using HiSeq Reagent v2.5 kit (Illumina), following manufacturer’s protocol.

### Enzyme-linked Immunosorbent Assays (ELISA)

PGLYRP-1 and sIL6Rα were assessed using Human PGLYRP1/PGRP-S DuoSet Elisa and Human IL-6 R alpha DuoSet Elisa (R&D Systems). Serum was diluted with 1% BSA in PBS (pH 7.2-7.4, 0.2 micron filtered) at 1:100 for PGLYRP-1 and 1:300 for sIL6Rα. ELISAs were performed according to manufacturer’s protocol. All samples were run in duplicate. Optical Densities were assessed at 450 and 540 using a Spectra Max M5 (Molecular Devices). The intra- and inter-plater CVs for PGLYRP-1 were 2.4% and 11.0%. The intra- and inter-plater CVs for sIL6Rα were 3.8% and 18.9%.

### Statistical Analysis

All statistical analyses were performed in *R* (https://www.r-project.org/) unless otherwise specified.

### Meta-Analysis

To identify datasets studies used in the meta-analysis, NCBI’s Gene Expression Omnibus (https://www.ncbi.nlm.nih.gov/geo) was searched using the search “(cord blood) AND “Homo sapiens”[porgn:_txid9606]” and was limited to study types that included expression profiling by array. This search yielded 352 studies that were further examined for cell types assessed and metadata reported. 17 studies met inclusion criteria of reporting metadata regarding at least one perinatal risk factor (e.g. gestational age at birth, newborn sex, birthweight, maternal pre-pregnancy BMI, smoke exposure, mode of delivery) and expression data from either whole cord blood or cord blood mononuclear cells derived from human subjects. Expression, feature, and subject demographic data was extracted using *geoquery* (61). If expression data was non-normalized, it was quantile normalized and log2 transformed. Six studies were excluded due to no variability in demographic data (i.e. only males) or low data quality, leaving 605 unique cord blood gene expression samples. To assess associations between gene expression and perinatal risk factors, univariate, inverse variance weighted, random effects models were constructed for genes using the *GeneMeta* package (62). Newborn sex (Male vs. Female), maternal pre-pregnancy BMI (continuous: 0= BMI<18.5, 1= 18.5≤BMI<25, 2= 25≤BMI<30, 3= 30≤BMI), gestational age at birth in weeks (continuous), and birthweight in grams (continuous) were assessed as perinatal risk factors. Significant genes were defined as Benjamini-Hochberg correct p-value < 0.01. The z-score for each gene was averaged across each univariate test (**Equation 1**) and termed pooled z-score. It was used to assess how likely a gene is effected by multiple risk factors. If a gene was not assessed in one of the univariate analyses while assessed in others, the missing data was inferred as a z-score of zero. To determine cell enrichment, genes expression in each cell as defined by HPA (34, 35) were modeled as a function of the pooled z-score using general additive model with cubic splines function. To determine biological processes enriched in the low vs. high risk individuals, the pooled z-score was used a pre-ranked list for gene set enrichment analysis for GO biologic processes using *GSEA* (63). Significantly enriched GO terms were defined as Benjamini-Hochberg correct p-value < 0.01. Spearman’s correlations between pooled z-score, univariate z-scores, and individual data set z-scores were determined in *R*.

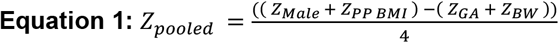

### RNA Sequencing Statistical Analysis (UIH Cohort)

The sequences were quality controlled by filtering out all low-quality reads (<25 on Phred quality score) and short reads (<50 bp). Transcripts were annotated using *salmon v0*.*12*.*0* and *Ensembl Homo sapiens* Genome Assembly GRCh38.12 (64). Transcript counts were aggregated in gene level counts using the *tximport* package in R (65). Genes with median counts across samples < 10 were filtered out, leaving 14,055 genes remaining whose expression normalized using median sum scaling. Normalized gene expression was modeled as function of the number of perinatal risk factors (gestational age < 37 weeks, birthweight < 3000 g, PP BMI >30, Male) using *DESeq2 (66)*. Genes were ranked for replication by their product of their pooled z-score and RNAseq z-score, termed replication score (RS) (**Equation 2**). A cutoff of RS > 3 was used to determine candidate genes associated with pediatric asthma risk. Candidate gene biologic process and cellular component enrichment was performed using STRING with default settings (67). Data from RNAseq is available on NCBI SRA database ID PRJNA577955.

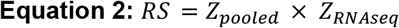

### Secondary Statistical Analysis of Mass Cytometry and ProSeek Data (Olin et al)

Methods for data acquisition for cell population percentages and protein abundances using mass cytometry and ProSeek are reported by Olin and colleagues (32). Cell population percentages were transformed using centered-log ratios. Cell populations and Protein abundances, and number of risk factors for each individual (gestational age < 37 weeks, male, birthweight < 3000g, birth via c-section) were correlated (Pearson’s method). For cell population correlations with number of risk factors, significance was defined as Bonferroni corrected p-value < 0.05. Proteins determined to neutrophil associated was determined by those expressed in CBMC (UIH cohort) and enriched in neutrophils HPA (34).

### Asthma and Pulmonary Outcomes Statistical Analysis (Project Viva)

Outcomes were assessed in Project Viva Categorical outcomes were modeled using logistic regression. Continuous outcomes were modeled using linear regression. Four regression models were used for each outcome: univariate/unadjusted, model1 adjusted for child’s demographics (gestational age at birth in weeks, birthweight adjusted for gestational age and sex, mode of delivery, sex, and race/ethnicity), model 2 adjusted for maternal demographics (PP BMI, race/ethnicity, level of education, atopy, antibiotic use during pregnancy, and smoking during pregnancy, 6 months, or 1 year), model 3 adjusted for both mother and child’s demographics (excluding maternal race/ethnicity). For regression models, PGLYRP-1 and sIL6Rα was log10 transformed and standardized to internal z-score. Subset analysis was performed by splitting the full data set by categorical variables and modeling outcomes as function of PGLYRP-1 in each subset using the univariate model. R^2^ for each variable in linear regression model 3 was determined using the *relaimpo* package (68). Mcfadden’s Pseudo-R^2^ for each variable in model 3 was calculated for logistic regression using full models minus the variable of interest.

## Data Availability

Data from RNAseq is available on NCBI SRA database ID PRJNA577955.

## Acknowledgments

Thanks to Kelly Liesse MD, Yishin Chang MS, and Wangfei Wang MS for their helpful comments regarding this manuscript.

## Supplemental Information

### Supplemental Data Description

**Title:** Supplemental Data 1

**Description:** Meta-Analysis of CBMC Gene Expression Associated with Newborn Sex

**Title:** Supplemental Data 2

**Description:** Meta-Analysis of CBMC Gene Expression Associated with Gestational Age

**Title:** Supplemental Data 3

**Description:** Meta-Analysis of CBMC Gene Expression Associated with Birthweight

**Title:** Supplemental Data 4

**Description:** Meta-Analysis of CBMC Gene Expression Associated with Maternal Pre-pregnancy BMI

**Title:** Supplemental Data 5

**Description:** Demographics of UIH Cohort

**Title:** Supplemental Data 6

**Description:** Result of mRNAseq UIH Cohort Modeling Gene Expression as a Function of Number of Risk Factors

**Title:** Supplemental Data 7

**Description:** Demographics of Project Viva Subset Dichotomized by Asthma Outcomes

---

**Figure S1.**
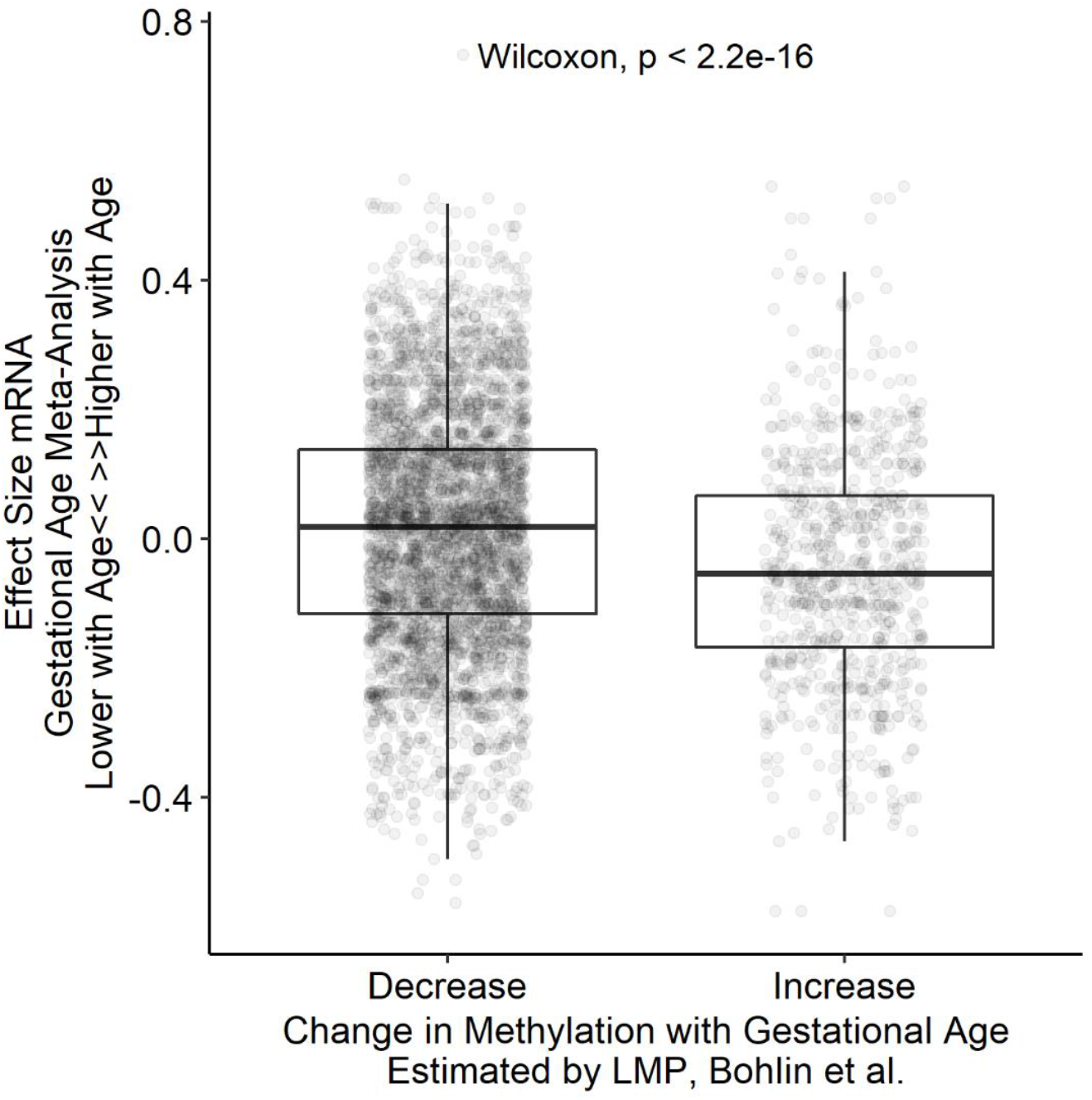
Association between Differentially Methylated Genes and Gene Expression Changes with Gestational Age. Comparison of effect size associated with gestational age for genes that were reported as differentially methylated by Bolin et al. (33). Gene with increased methylation associated with gestational age demonstrate reduced expression with increasing gestational age.

**Figure S2.**
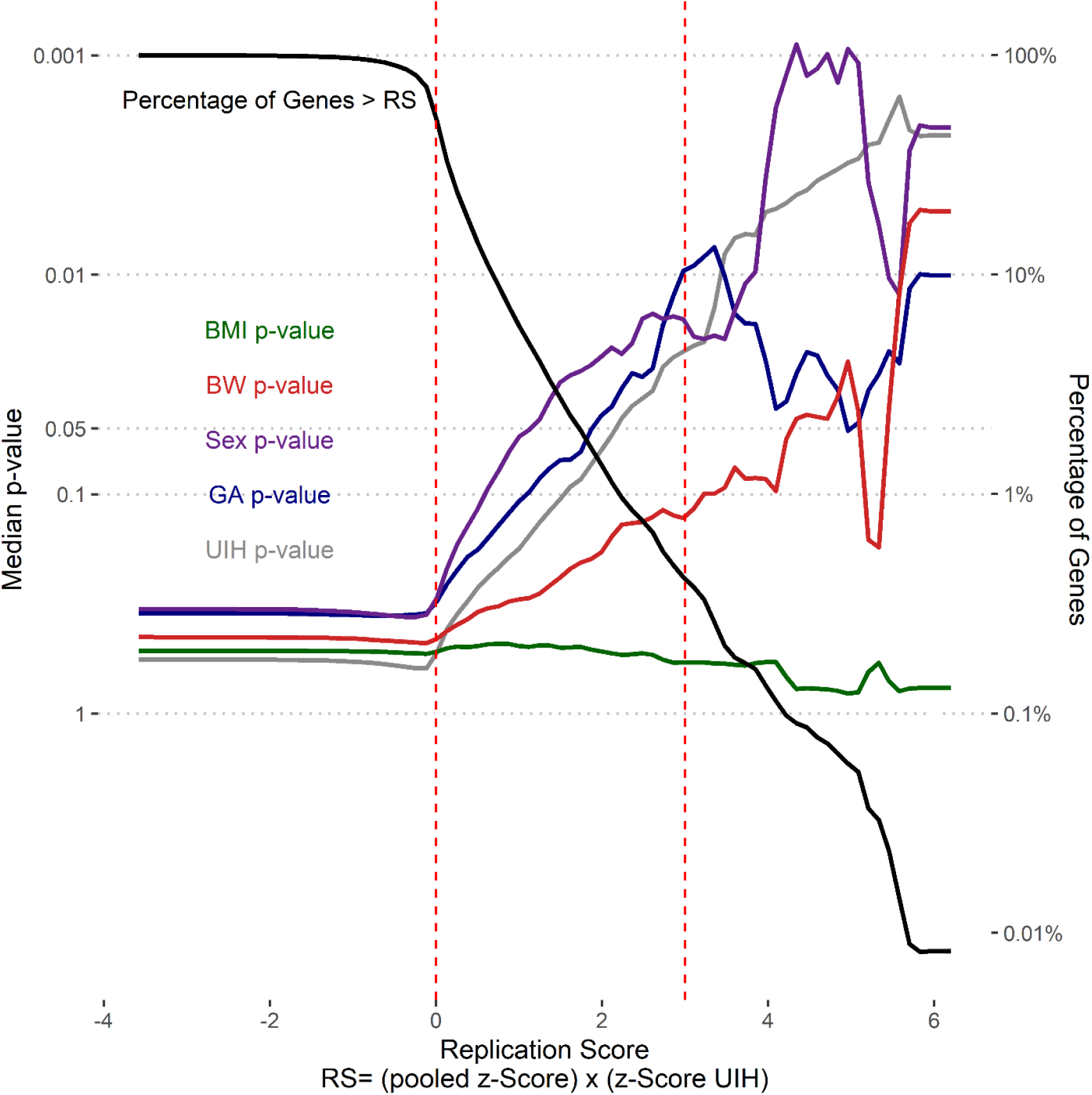
Replication Score Enriches for Genes Associated with Multiple Risk Factors. Splines (colored according to analysis) of median p-values (left y-axis) for genes with replication scores greater than corresponding cut-off (x-axis). Percentage of genes with replication score greater than corresponding cut-off. Vertical dashed lines two cutoffs: RS > 0 and RS >3.

**Figure S3.**
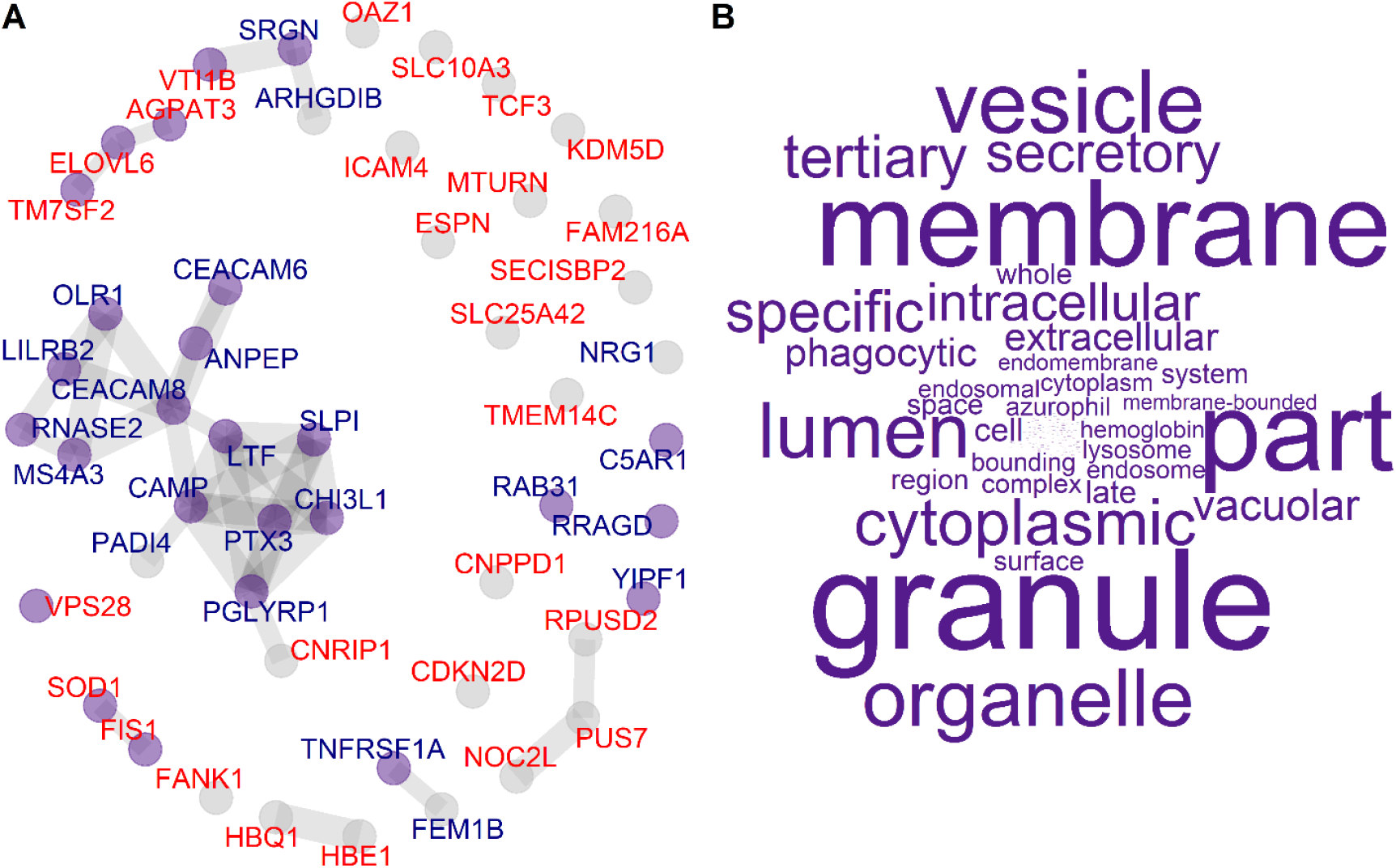
Protein-Protein Interaction Network of Candidate Genes. **(A)** Protein-Protein interaction network of candidate genes inferred from STRING (67). Nodes are labeled by risk association: low (blue) and high (red) risk candidate genes. Nodes are colored (purple) if they are associated with GO cellular component term enrichment. **(B)** Word clouds of GO terms significantly enriched in candidate genes.

**Figure S4.**
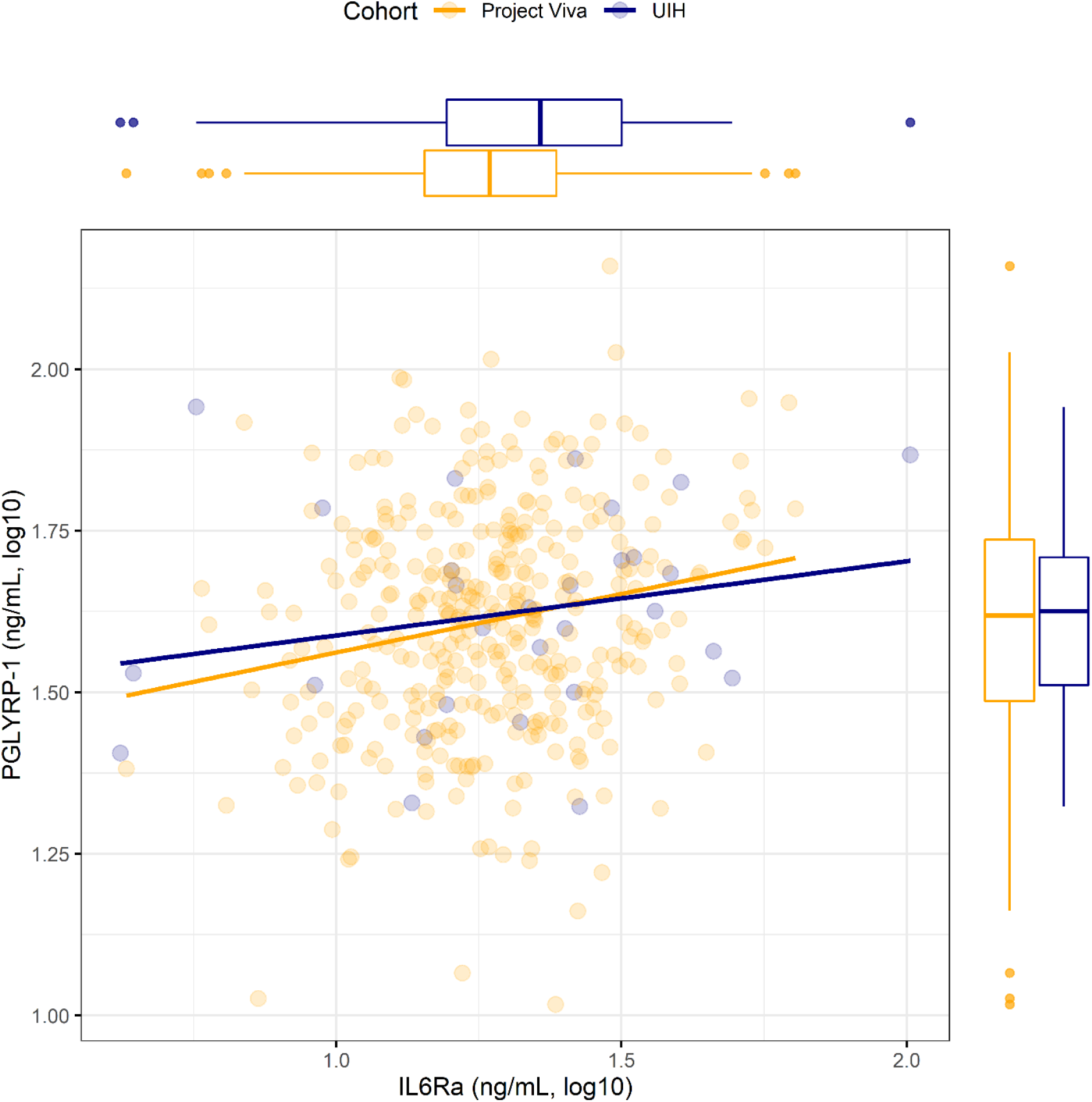
Association between PGLYRP-1 and sIL6Rα in UIH and Project Viva Cohorts. Scatter plot displaying association between PGLYRP-1 and sIL6Rα in UIH (blue) and Project Viva (yellow) cohorts. Univariate regression lines are shown for both cohorts. Distributions for PGLYRP-1 and sIL6Rα are shown in the margins for each cohort.

**Figure S5.**
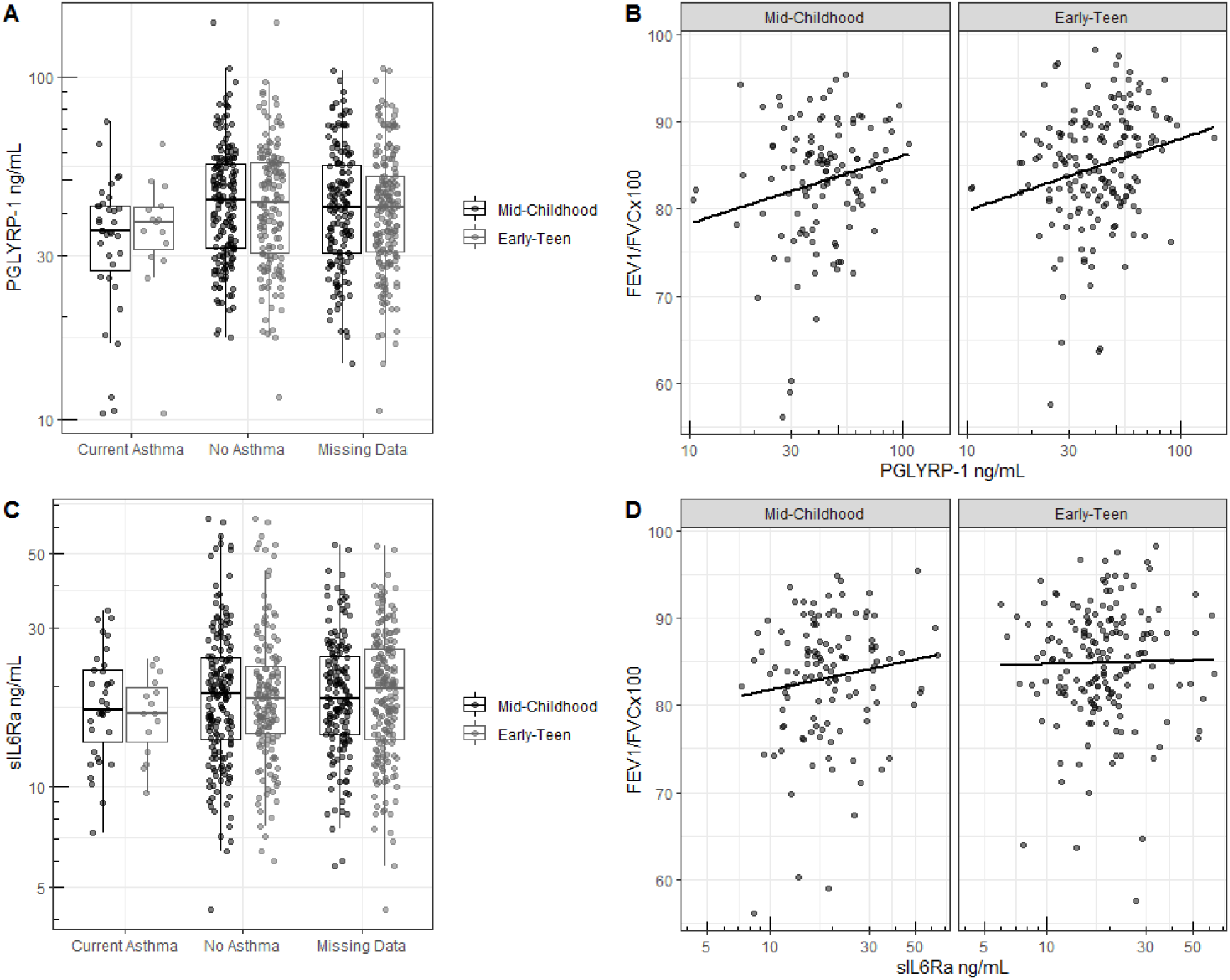
Cord Blood Serum Proteins in Relationship to Outcomes. **(A)** PGLYRP-1 concentration in umbilical cord blood serum in relationship to current asthma determined by questionnaire response and **(B)** FEV1/FVC×100 at mid-childhood and early-teenage follow-ups. sIL6Rα concentration in umbilical cord blood serum in relationship to current asthma determined by questionnaire response and **(D)** FEV1/FVC×100 at mid-childhood and early-teenage follow-ups.

**Figure S6.**
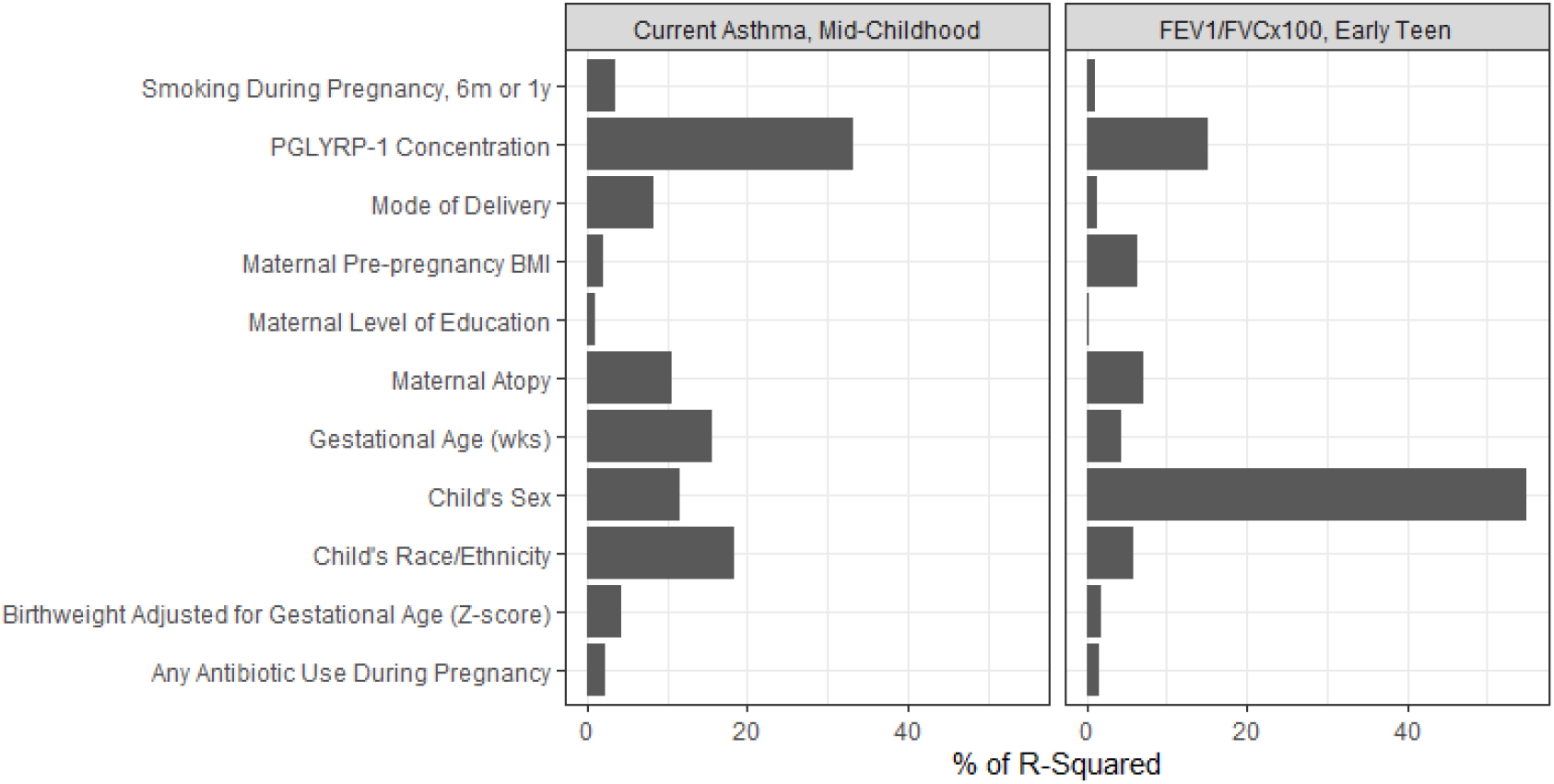
Relative importance of predictors for pediatric asthma and FEV1/FVC. Relative importance, displayed as percent of variance explained, for variables used in regressions (Table 3, Model 3) for current asthma at mid-childhood and FEV1/FVC in early teen years. Variance estimated for logistic regression as Mcfadden’s Pseudo-R^2^.

**Figure S7.**
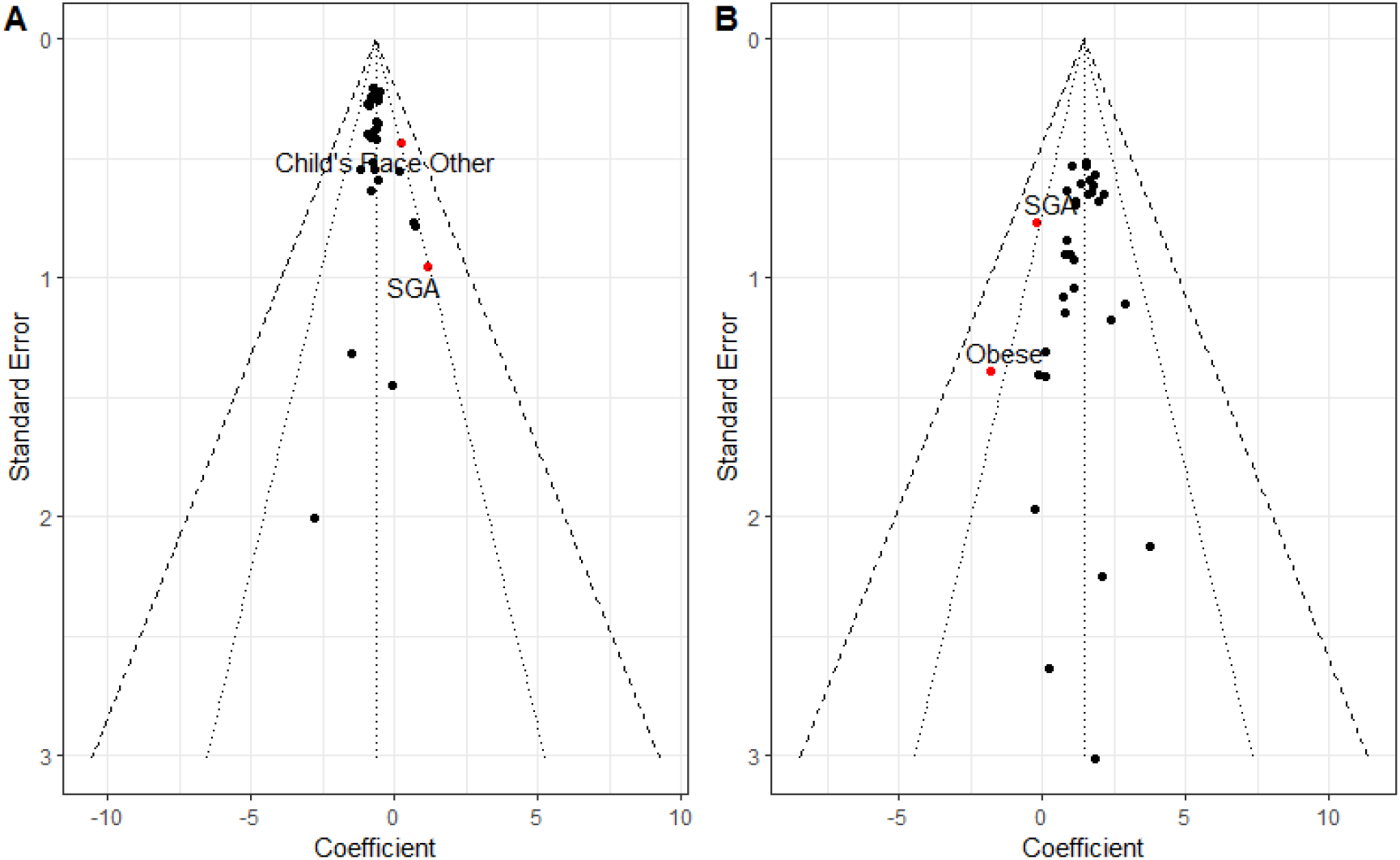
Subset analysis for all covariates used in regression models. Funnel plot demonstrating relationship effect size estimates and measurement error for subset analyses for **(A)** current mid-childhood asthma and **(B)** FEV1/FVC×100 in early teen years. 95% CI (botted lines) and 99% CI (dashed lines) displayed.

## Notes

**Competing Interests:** The authors declare that they have no relevant conflicts of interest.

### Competing Interest Statement

The authors have declared no competing interest.

### Clinical Trial

NCT02820402

### Funding Statement

This work was funded by the National Institutes of Health, grant nos. R01 AI 053878,
F30 HL 136001, R01 HD 034568, and UH3 OD 023286, the Robert Wood Johnson Foundation Nurse Faculty Scholars Program 72117, the College of Nursing Deans Award, and University of Illinois at Chicago Department of Medicine.

### Author Declarations

The current study was approved by the University of Illinois at Chicago IRB, 20160326 and 20150353, and the IRB of Harvard Pilgrim Health Care.

